# Development of a COVID-19 Vaccination Anxiety Scale to measure COVID-19 vaccine anxiety in Japanese adults

**DOI:** 10.1101/2025.09.03.25332753

**Authors:** Time Fukuda, Ranno Haruyama, Yuto Tanaka, Shuma Natori, Kohei Koiwa, Koubun Wakashima

## Abstract

Since 2020, COVID-19 vaccines have been rapidly developed and distributed free of charge in several countries, but there remains deep-seated distrust and anxiety regarding their efficacy and safety. This study aimed to develop the C19-VAS to measure COVID-19 vaccine anxiety in Japanese adults. 500 Japanese participants were recruited through the web survey service. To assess test-retest reliability, the same participants were surveyed again after a four-week interval. These participants responded to the C19-VAS, the Japanese version of the Fear of COVID-19 Scale (FCV-19S), the Japanese version of the 7C of Vaccination Readiness (7C), the Japanese version of the Generic Conspiracist Beliefs Scale (GCBS-J), the Japanese versions of the Perceived Vulnerability to Disease (PVD-J), and socio-demographic variables. Results indicated the factor structure of the C19-VAS as including 25 items and 3 factors: “Anxiety Related to Vaccine Confidence (α = .96, ω = .96),” “Emotional Symptoms of Anxiety (α = .91, ω = .91),” and “Anxiety about Infection after Vaccination (α = .76, ω = .78).” The confirmatory factor analysis findings supported the 3 factors structure of the C19-VAS across gender with good fit statistics: CFI = .868, RMSEA = .071, SRMR = .079, AIC = 1434.177. The scale showed adequate construct validity, as indicated by significantly correlations with the FCV-19S-J (r = .44), 7C (r = −.59), GCBS-J (r = .55) and PVD-J (r = .19). Additionally, the ICC between Time 1 and Time 2 for “Anxiety Related to Vaccine Confidence” was .94, “Emotional Symptoms of Anxiety” was .91, and “Anxiety about Infection after Vaccination” was .81. This study clarified the factor structure of the C19-VAS and demonstrated its good internal consistency and test-retest reliability.

## Introduction

The global spread of COVID-19 has served as a catalyst for society to recognize the importance of vaccine development and distribution. Since 2020, COVID-19 vaccines have been rapidly developed and distributed free of charge in several countries, playing a central role in public health policies. However, despite the importance and prevalence of vaccines, there remains deep-seated distrust and anxiety regarding their efficacy and safety [1]. Consequently, the challenge of convincing individuals who hesitate to be vaccinated has become a common global issue.

Despite the persistence of vaccine hesitancy or anxiety, scientific evidence confirms the effectiveness of COVID-19 vaccines in preventing severe illness. For example, a study in the United States reported that the Omicron-variant-specific vaccine (BNT162b2 XBB) had a 62% efficacy in preventing hospitalization and a 73.3% efficacy in preventing emergency department visits [2]. Domestic research in Japan has also shown that while the vaccine has limited effectiveness in preventing infection, it has an additional effect in preventing hospitalization. [3]. Furthermore, the effectiveness of the COVID-19 vaccine may weaken over time, and regular vaccination is recommended [4]. Despite the proven effectiveness of the vaccine, psychological factors, such as anxiety and distrust, are cited as reasons for the lack of enthusiasm for vaccination. Lazarus et al. [5] reported that in an international comparative survey of 13,426 people in 19 countries, 71.5% of respondents said they would like to be vaccinated if the vaccine was safe and effective, but there were significant differences in acceptance rates between countries. Asian countries, such as China and South Korea, showed high levels of vaccine acceptance, with rates exceeding 80%, while some European and American countries remained at 50–60%. Such differences are thought to be influenced by not only individual rational judgments but also social factors such as cultural background, trust in the government, and differences in healthcare systems. Japan was not included in this survey, but according to a domestic survey by Fukunaga [6], the main reason individuals hesitate to get vaccinated is “concerns about safety.” These concerns involve individual health awareness, distrust of pharmaceutical companies and the government, and conspiracy theories, making them complex and not easily categorized as a simple yes-or-no issue.

Anxiety about vaccination is not unique to COVID-19; it has also been observed in the past with the introduction of influenza and HPV vaccines. However, vaccine-related anxiety is often discussed within the framework of underestimating scientific risks and overestimating emotional risks. The intensity and content of such anxiety can vary significantly depending on the social context [7]. The factors specific to COVID-19 vaccine anxiety include, first, the rapid spread and confusion of information in an unprecedented pandemic; second, unfounded anxiety about the new mRNA technology; and third, the inclusion of altruistic motives such as “for the sake of society” and “for the sake of others” in vaccination intentions. These factors are believed to have caused complex and multifaceted psychological reactions. Romate et al. [8] conducted a systematic review and identified fear of infection and severe illness, concerns about vaccine safety and side effects, and anxiety about interactions with preexisting conditions as the main factors associated with COVID-19 vaccine hesitancy. Additionally, trust in vaccines in general, trust in the government and healthcare institutions, beliefs in conspiracy theories, and emotional factors such as health anxiety also significantly influence vaccination anxiety. Given the complex interplay among these factors, it is necessary to develop a scale that considers these multifaceted components to accurately assess COVID-19 vaccination anxiety.

Various scales were developed to capture this multifaceted structure. For example, the Oxford COVID-19 Vaccine Hesitancy Scale developed by Freeman et al. [9] succinctly captures vaccination attitudes and refusal tendencies but does not sufficiently address the content of anxiety. Additionally, its validity has only been verified in relation to the fear of infection, leaving room for further validation. The COVID-19 Vaccine Concerns Scale by Gregory et al. [10] captures concerns specific to COVID-19 but does not address anxiety or fear itself. Therefore, it has a limited ability to capture the diversity of psychological backgrounds. Furthermore, the Multidimensional Vaccine Hesitancy Scale by Balgiu et al. [11] takes a comprehensive approach with eight factors; however, some of these factors are not specific to COVID-19 and instead focus on elements common to general vaccination (cost, inconvenience of vaccination, pain, etc.), limiting its application to COVID-19. Furthermore, there is no standardized scale in Japan, highlighting the need to develop measurement tools tailored to the characteristics of the Japanese population.

Based on the above background, this study aimed to develop the COVID-19 Vaccination Anxiety Scale (C19-VAS) to measure COVID-19 vaccine anxiety in Japanese adults from multiple perspectives. To examine the construct validity of the C19-VAS, we verified its relationship with the following four existing scales:

The first measure was the Japanese version of the Fear of COVID-19 Scale (FCV-19S-J) [12], which assesses the fear of COVID-19 infection. Individuals with higher levels of fear of the COVID-19 infection are more sensitive to vaccine side effects and efficacy, paradoxically increasing their anxiety toward vaccination. Thus, FCV-19S-J and C19-VAS were expected to show a positive correlation.

The second is the Japanese version of the 7C of Vaccination Readiness (7C) [13], which assesses the predictive factors for vaccination intent. As those with higher anxiety about COVID-19 vaccines are likely to have lower readiness for vaccination, it was predicted that the 7C and C19-VAS scores will show a negative correlation.

The third is the Japanese version of the Generic Conspiracist Beliefs Scale (GCBS-J) [14], which assesses general tendencies toward conspiracist beliefs. Individuals with strong conspiratorial thinking tendencies are more likely to harbor distrust regarding the safety of the COVID-19 vaccine and the government’s intentions, leading to heightened vaccination anxiety. Therefore, it was predicted that the GCBS-J and C19-VAS scores would show a positive correlation.

The fourth was the Japanese version of the Perceived Vulnerability to Disease Scale (PVD-J) [15], which assesses awareness of vulnerability to infectious diseases. Individuals with a high perception of vulnerability to infectious diseases are likely to be sensitive to the health effects of vaccines; therefore, it was predicted that the PVD-J and C19-VAS will show a positive correlation.

## Material and Methods

### Participants and procedure

The survey, conducted between April and May 2025, targeted 500 participants aged 18 and over through the web survey service (Freeasy). Of the 500 participants, we excluded 3 with incomplete responses, 5 physicians or medical professionals, and 72 who provided inappropriate responses to the Instructional Manipulation Check task [16], leaving 420 participants (208 men and 212 women; *M* = 60.33, *SD* = 10.7) for analysis. Additionally, to assess test-retest reliability, the same participants were surveyed again after a four-week interval. Among them, 356 participants who completed both surveys and answered correctly on the Instructional Manipulation Check task [16] were included in the analysis. Table 1 presents the participants’ characteristics.

**Table 1.** Demographic Characteristics of Participants.

|  | *N* | (%) | *Mean* | (*SD*) |
| --- | --- | --- | --- | --- |
| Age | 420 |  | 60.3 | (10.7) |
| Gender |  |  |  |  |
| Male | 208 |  |  |  |
| Female | 212 |  |  |  |
| Previous COVID-19 Infection |  |  |  |  |
| Yes | 119 | (28.3) |  |  |
| No | 301 | (71.7) |  |  |
| Chronic diseases |  |  |  |  |
| Yes | 104 | (24.8) |  |  |
| No | 316 | (75.2) |  |  |
| Living with Family |  |  |  |  |
| Yes | 339 | (80.7) |  |  |
| No | 81 | (19.3) |  |  |
| Number of Family Members Living Together |  |  |  |  |
| 1 | 98 | (23.3) |  |  |
| 2 | 119 | (28.3) |  |  |
| 3 | 82 | (19.5) |  |  |
| 4 | 32 | (7.6) |  |  |
| 5 | 4 | (1.0) |  |  |
| 6 | 3 | (0.7) |  |  |
| 7 | 1 | (0.2) |  |  |
| Presence or Absence of Children |  |  |  |  |
| Presence | 190 | (45.2) |  |  |
| Absence | 230 | (54.8) |  |  |

### Ethical consideration

After the survey’s purpose was presented, the form stated that participation was voluntary and anonymous. Personal information would not be disclosed to third parties. Only those who agreed to cooperate in the survey would be able to proceed to the questionnaire. The Tohoku University Graduate School of Education’s Ethics Committee approved this study (ID: 24-1-092).

### Measures

The same questions were asked in the first and second surveys.

#### Sociodemographic variables

Respondents were asked to provide information on their age, gender, pregnancy status, occupation, history of COVID-19 infection, presence of chronic diseases, sources of information on COVID-19, presence of family members living in the same household, and number of family members living in the same household.

#### C19-VAS

The scale developed by Al Baseer and Shaheen [17] was translated into Japanese and slightly modified with permission from the original authors. While Al Baseer and Shaheen [17] used a 3-point scale, a 4-point to 6-point scale was considered the optimal range [18]. Using a 3-point scale or lower may reduce reliability and validity [19]. Therefore, in this study, participants were asked to respond to each item using a 5-point scale ranging from “1 = does not apply” to “5 = applies.” Higher scores indicated greater anxiety about the COVID-19 vaccination. Additionally, when translating and revising the items, we confirmed the content validity with one faculty member with qualifications in clinical psychology and two graduate students specializing in clinical psychology.

#### FCV-19S-J

The FCV-19S-J [12] was used. This scale consists of 7 items, with responses ranging from “1 = Not at all” to “7 = Strongly agree” on a 7-point scale. In this study, the total score was used for analysis.

#### 7C

The 7C [13] was used. This scale consists of 21 items, with 3 items for each of the 7 factors, and responses were acquired through a 7-point scale ranging from “1 = Strongly disagree” to “7 = Strongly agree.” In this study, the total score was used for analysis.

#### GCBS-J

The GCBS-J [14] was used. This scale consists of 15 items across 2 factors, with responses ranging from “1 = Definitely incorrect” to “5 = Definitely correct” on a 5-point scale. In this study, the total score was used for analysis.

#### PVD-J

The PVD-J [15] was used. This scale consists of 15 items across 2 factors, with responses ranging from “1 = Not at all applicable” to “7 = Very applicable” on a 7-point scale. In this study, the total score was used for analysis.

### Data analysis

Confirmatory factor analysis (CFA) and multiple-group CFA were performed using IBM SPSS AMOS 29. All other analyses were conducted using the IBM SPSS Statistics version 29.

When translating into Japanese, we shifted from a 3-point scale to a 5-point scale and revised some of the items. Consequently, we decided to conduct an exploratory factor analysis (EFA). The data were randomly sampled into two groups: training and testing. First, an EFA was performed on the training data (N = 210) using maximum likelihood estimation and Promax rotation, excluding unnecessary items. CFA was performed on the test data (N = 210). As an additional analysis, multiple-group CFA was conducted to examine gender invariance. To test goodness of fit, we conducted the following analyses: comparative fit index (CFI), root mean square error of approximation (RMSEA), standardized root mean square residual (SRMR), and Akaike information criterion (AIC). The cutoff values for acceptable model fit used in this study were as follows: RMSEA < .10 for acceptable fit and < .06 for good fit; CFI > .90 for acceptable fit and > .95 for good fit; and SRMR < .10 for acceptable fit and < .08 for good fit. Error correlation was assumed to be related to the modified index (MI) if the goodness of fit was inadequate. Reliability was calculated for Cronbach’s alpha coefficients (α) and McDonald’s omega coefficients (ω). Correlations between the C19-VAS score and other measures were established by calculating Pearson’s correlation coefficients. The test-retest correlation coefficient was calculated using ICC (intraclass correlation coefficient, two-way random effects, and absolute agreement).

## Results

### Factor validity

As no items showing ceiling or floor effects were identified, factor analysis was performed on the 30 items of the original C19-VAS. The initial eigenvalues include 14.08, 2.85, 1.75, 1.17, 0.99, in order from the first factor. Based on the scree plot criterion and interpretability of the factors, a three-factor structure was deemed appropriate.

Next, factor analysis was performed, assuming 3 factors; 4 items with factor loadings below 0.40 and 1 item showing double loading were sequentially deleted. The final pattern matrix and inter-factor correlations are shown in Table 2. The cumulative factor contribution rate was 67.59% for the 3 factors across the 25 items. The first factor, “Anxiety Related to Vaccine Confidence,” consists of items such as “I lose confidence in the vaccine’s effectiveness owing to its variety and the different samples tested” and “I am worried about the side effects of the vaccine.” The second factor, “Emotional Symptoms of Anxiety,” comprises items such as “I am overwhelmed by feelings of fear due to excessive thinking about illness and death after vaccination” and “I tremble at the mere thought of vaccinating my family.” The third factor, “Anxiety about Infection after Vaccination,” includes items such as “Even after vaccination, I still dread the thought of being in contact with large numbers of people at work or school” and “I fear attending social gatherings despite being vaccinated.”

**Table 2.**
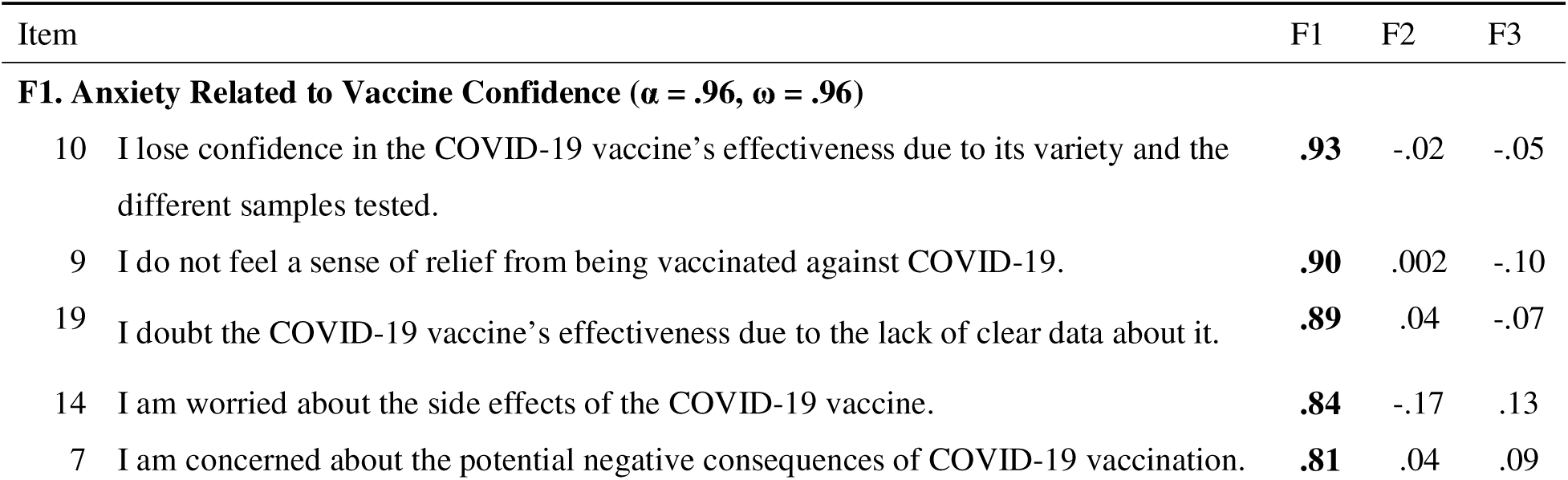
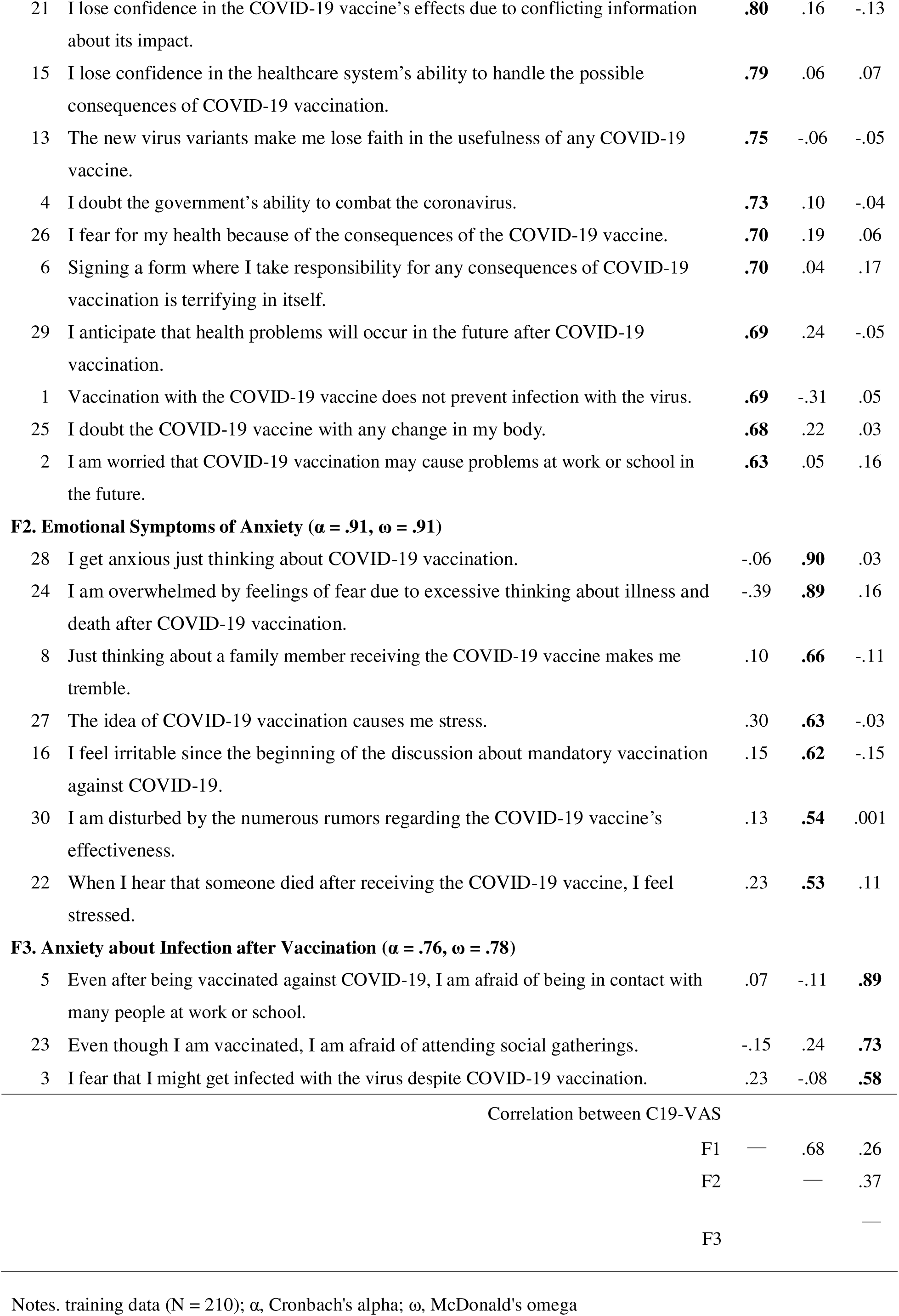
Exploratory Factor Analysis of the C19-VAS.

Finally, a CFA was performed using the three factors obtained through EFA as latent variables. The results of the CFA of the 3-factor model, with paths drawn from each latent variable to the corresponding items, showed that the model fit indices were χ2 = 781.311, p < .001, df = 272, CFI = .894, RMSEA = .095 [90% CI = .087–.102], SRMR = .073, AIC = 887.311, indicating insufficient model fit. MI was used to improve the model fit. The MI between items 6 and 7 (MI = 28.94) was the highest. Therefore, a within-factor error covariance between items 8 and 11 was included, and the model was modified. The results indicated that the modified model was more acceptable (χ2 = 750.866, p < .001, df = 271, CFI = .901, RMSEA = .092 [90% CI = .084–.100], SRMR = .072, AIC = 858.866) and was used as the final model. Descriptive statistics of the C19-VAS and other scales are presented in Table 3.

**Table 3.** Descriptive Statistics of the Sample’s Responses to the Study Scales.

| Variable | Mean | SD | Minimum | Maximum |
| --- | --- | --- | --- | --- |
| **C19-VAS** | 74.85 | 19.74 | 25 | 125 |
| Anxiety Related to Vaccine Confidence ($\alpha = .96$, $\omega = .96$) | 48.32 | 13.32 | 15 | 75 |
| Emotional Symptoms of Anxiety ($\alpha = .91$, $\omega = .91$) | 17.59 | 6.18 | 7 | 35 |
| Anxiety about Infection after Vaccination ($\alpha = .76$, $\omega = .78$) | 8.94 | 2.76 | 3 | 15 |
| **FCV-19S-J ($\alpha = .90$, $\omega = .90$)** | 15.45 | 5.09 | 7 | 29 |
| **7C** | 84.12 | 14.33 | 33 | 131 |
| Confidence ($\alpha = .79$, $\omega = .80$) | 12.00 | 3.27 | 3 | 21 |
| Complacency ($\alpha = .86$, $\omega = .86$) | 13.54 | 3.53 | 3 | 21 |
| Constraints ($\alpha = .73$, $\omega = .74$) | 12.30 | 3.09 | 3 | 21 |
| Calculation ($\alpha = .44$, $\omega = .47$) | 10.24 | 2.42 | 3 | 21 |
| Collective Responsibility ($\alpha = .88$, $\omega = .90$) | 13.12 | 3.45 | 3 | 21 |
| Compliance ($\alpha = .75$, $\omega = .76$) | 10.44 | 3.06 | 3 | 20 |
| Conspiracy ($\alpha = .76$, $\omega = .76$) | 12.47 | 2.86 | 3 | 21 |
| **GCBC-J** | 37.17 | 12.85 | 15 | 75 |
| General Conspiracist Beliefs ($\alpha = .96$, $\omega = .96$) | 30.19 | 10.30 | 12 | 60 |
| Extraterrestrial Conspiracist Beliefs ($\alpha = .92$, $\omega = .92$) | 6.97 | 2.94 | 3 | 15 |
| **PVD-J** | 62.67 | 10.59 | 27 | 93 |
| Perceived Infectability ($\alpha = .84$, $\omega = .83$) | 26.54 | 5.99 | 7 | 48 |
| Germ Aversion ($\alpha = .78$, $\omega = .78$) | 36.13 | 7.11 | 11 | 56 |

Notes. $\alpha$ , Cronbach's alpha; $\omega$ , McDonald's omega

### Multiple-group CFA

To examine whether the same factor patterns apply to the male and female subsamples, we examined the measurement invariance by gender. Specifically, we confirmed the fit for each gender, then compared four models across datasets: a configuration-invariant model with no equivalence constraints on each dataset; a weak measurement invariance model with equivalence constraints only on factor loadings; a strong measurement invariance model with equivalence constraints on both factor loadings and intercepts; and a strict measurement invariance model with equivalence constraints on both factor loadings, intercepts, and error variables.

Regarding the goodness of fit for each gender, male individuals had χ2 = 559.142, p < .001, df = 271, CFI = .891, RMSEA = .104 [90% CI = .091–.116], SRMR = .080, AIC = 667.142, and female individuals had χ2 = 589.683, p < .001, df = 271, CFI = .859, RMSEA = .104 [90% CI = .092–.115], SRMR = 0.085, AIC = 697.683. Although the CFI criterion was not met, the RMSEA was slightly above the criterion value and the SRMR was below the criterion value for both men and women, indicating an acceptable level of fit. We conducted a multi-population simultaneous analysis by gender for the configuration- and measurement-invariant models. The results showed that the fit of the configuration-invariant model was χ2 = 1148.837, p < .001, df = 542, CFI = .877, RMSEA = .073 [90% CI = .067–.079], SRMR = .080, AIC = 1464.837. The fit of the weak measurement invariance model was χ2 = 1186.602, p < .001, df = 563, CFI = .873, RMSEA = .073 [90% CI = .067–.079], SRMR = .076, AIC = 1460.602. The fit of the strong measurement invariance model was χ2 = 1220.149, p < .001, df = 588, CFI = .871, RMSEA = .072 [90% CI = .066–.078], SRMR = .076, AIC = 1444.149. Finally, the fit of the strict measurement invariance model was χ2 = 1260.177, p < .001, df = 613, CFI = .868, RMSEA = .071 [90% CI = .066–.077], SRMR = .079, AIC = 1434.177. Although the CFI value decreased as the number of equivalence constraints increased, the maximum decrease was only .009, and the RMSEA and SRMR values indicated a good fit. Furthermore, when comparing each model, including the AIC values, the strict measurement invariance model showed the best values among the four models, confirming strict measurement invariance (Table 4). Based on the above, as strict measurement invariance was confirmed for gender, the same three-factor structure was applied to construct the scale regardless of gender.

**Table 4.** Measurement Invariance of Model among Gender Groups for C19-VAS.

| | $\chi^2$ | df | CFI | RMSEA | SRMR | AIC |
| --- | --- | --- | --- | --- | --- | --- |
| Configural model | 1148.837 | 542 | .877 | .073 | .080 | 1464.837 |
| Metric invariance model | 1186.602 | 563 | .873 | .073 | .076 | 1460.602 |
| Scalar invariance model | 1220.149 | 588 | .871 | .072 | .076 | 1444.149 |
| Strict invariance model | 1260.177 | 613 | .868 | .071 | .079 | 1434.177 |

Notes. test data (N = 210); CFI, comparative fit index; RMSEA, root mean square approximation; SRMR, standardized root mean square residual; and AIC, Akaike information criterion.

### Examination of gender differences

To examine gender differences in COVID-19 vaccine anxiety, a t-test (two-tailed) was conducted on 420 participants at Time 1. The results showed a significant gender difference (t (418) = 2.34, p = .02, d = .23). This indicates that women had higher total scores on the scale than men.

### Reliability

Cronbach’s α and McDonald’s ω were calculated for the subscales of C19-VAS using data from 420 participants at Time 1. The first factor had α = .96 and ω = .96; the second factor had α = .91 and ω = .91; and the third factor had α = .76 and ω = .78, demonstrating sufficient internal consistency. Additionally, the ICC between Time 1 and Time 2 for “Anxiety Related to Vaccine Confidence” was .94 (95% CI [.92–.96]), “Emotional Symptoms of Anxiety” was .91 (95% CI [.89–.93]), and “Anxiety about Infection after Vaccination” was .81 (95% CI [.77–.85]).

### Construct validity

To verify the construct validity of the C19-VAS, we performed a correlation analysis with the total scores of the FCV-19S-J, 7C, GCBS-J, and PVD-J at Time 1. When examining the correlation coefficients between the C19-VAS total score and FCV-19S-J, 7C, GCBS-J, and PVD-J, there was a moderate to strong positive correlation with FCV-19S-J (r = .44, p < .001), a strong negative correlation with 7C (r = −.59, p < .001), a strong positive correlation with GCBS-J (r = .55, p < .001), and a small to moderate positive correlation with PVD-J (r = .19, p < .001).

## Discussion

This study aimed to develop the C19-VAS to measure COVID-19 vaccine anxiety in Japanese adults. Factor analysis revealed that the C19-VAS was constructed with 3 factors and 25 items: “Anxiety Related to Vaccine Confidence,” “Emotional Symptoms of Anxiety,” and “Anxiety about Infection after Vaccination.” These three factors differ from those developed by Al Baseer and Shaheen [17]. This is likely owing to the change from a 3-point scale to a 5-point scale and some revisions to the scale items. The scale developed by Al Baseer and Shaheen [17] was designed for university faculty, whereas the C19-VAS was revised for application in the general adult population. Therefore, the factor structure assumed by Al Baseer and Shaheen [17] was not observed. In the CFA, assuming an error correlation between item 6 and item 7 indicated an acceptable fit. The results of the correlations between each item showed that these two items had a high degree of correlation with each other compared to the other items on the scale. These two items were considered to be different from other items in that they did not refer to specific contents of anxiety within the factor “Anxiety Related with Vaccine Confidence.”

A multiple-group CFA was conducted by gender. For both men and women, CFI fell below, RMSEA was slightly above, and SRMR was below the benchmark value, indicating an acceptable but not ideal fit. Furthermore, when comparing the gender-invariant, weak measurement invariance, strong measurement invariance, and strict measurement invariance models, all models showed good fit in terms of RMSEA and SRMR. Among the four models, strict measurement invariance showed the best values, including AIC values, confirming strict measurement invariance. These results indicated that the three-factor model developed in this study was applicable to both men and women.

Next, the results of the gender difference analysis showed that women had higher COVID-19 vaccination anxiety than men. The results were consistent with those of Sekizawa et al. [20], who showed that women tended to be negative about COVID-19 vaccination. Women experience more side effects from the COVID-19 vaccine owing to hormonal differences between men and women [21], which may have contributed to gender differences in anxiety about COVID-19 vaccination. Additionally, it has been pointed out that women generally have higher sensitivity to COVID-19-related risks [22], and such personality traits may also influence gender differences in anxiety.

Internal consistency and test-retest reliability were also examined. The α coefficient and ω coefficient in C19-VAS returned sufficient values. Test-retest reliability also showed sufficient values for all three factors, confirming test-retest reliability. Subsequently, to verify construct validity, a correlation analysis was conducted between the C19-VAS at Time 1 and the total scores on the FCV-19S-J, 7C, GCBS, and PVD-J. The results showed positive correlations with FCV-19S-J, GCBS, and PVD-J, and a negative correlation with 7C. The correlation coefficient with PVD-J was lower than those with other scales, which was likely due to differences in the factors of PVD-J, namely, “perceived infectability” and “germ aversion.” When the correlation between each factor and C19-VAS score was examined, a significant correlation (r = .20, p < .001) was confirmed with the latter but no correlation (r = .09, p = .06) was confirmed with the former. This suggests that perceived infectability is not associated with COVID-19 vaccine anxiety, whereas perceived discomfort in situations where pathogens are likely to attach is associated with COVID-19 vaccine anxiety. Duncan et al. [23] distinguished between “perceived infectability,” which is a response based on rational judgment, and “germ aversion,” which is a response based on intuitive judgment. Items such as “I don’t want to wear secondhand clothes because I don’t know who last wore them” reflect “germ aversion” and measure anxious tendencies toward infection risks that lack empirical basis [15]. Regarding the COVID-19 vaccine, providing unfounded conspiracy theories or misinformation increases hesitancy toward vaccination [24]. Many conspiracy theories and rumors exploit intuitive reactions rather than theoretical evidence of stroke anxiety. Therefore, COVID-19 vaccine anxiety and “germ aversion” share a similar cognitive tendency to avoid risks through immediate judgments based on weak evidence, which may explain the observed correlation. Meanwhile, “perceived infectability” is a response based on rational judgment [23]. Therefore, no significant correlation was demonstrated between “perceived infectability” and C19-VAS. Further investigation is necessary in this regard. Based on these results, the C19-VAS was considered to have sufficient reliability and validity.

This study has some limitations. First, healthcare workers were excluded from the analysis. Healthcare workers received COVID-19 vaccines earlier and more frequently than the general population. Therefore, the C19-VAS, which was developed for the general adult population, has a different structure; thus, developing a scale specifically for healthcare workers is necessary. Second, this study did not include questions about vaccination history before taking the COVID-19 vaccine. The presence or absence of vaccination experience may influence attitudes toward the vaccine and anxiety. In the future, it will be necessary to collect detailed information on vaccination history, such as the number of doses administered and the presence or absence of adverse reactions, and examine their association with vaccination anxiety.

## Conclusions

This study clarified the factor structure of the C19-VAS and demonstrated its good internal consistency and test-retest reliability. This scale will be useful for future epidemiological surveys, intervention studies, and public health policy planning. Furthermore, it can be used as a basic framework for assessing vaccination anxiety for not only COVID-19 but also other infectious disease outbreaks and when vaccines are introduced.

## Data Availability

All relevant data are within the Supporting Information files.

## Acknowledgements

We would like to express our sincere gratitude to all the participants who cooperated with this research. We would like to thank Editage for English language editing.

## References

1. Sallam M. COVID-19 Vaccine Hesitancy Worldwide: A Concise Systematic Review of Vaccine Acceptance Rates. Vaccines. 2021;9(2):160. doi: 10.3390/vaccines9020160

2. Tartof SY, Slezak JM, Frankland TB, Puzniak L, Hong V, Ackerson BK, et al. Estimated Effectiveness of the BNT162b2 XBB Vaccine Against COVID-19. JAMA Internal Medicine. 2024;184(8):932–940. doi: 10.1001/jamainternmed.2024.1640. PMID: 38913355; PMCID: PMC11197441.

3. Institute of Tropical Medicine, Nagasaki University. Vaccine Effectiveness Real-Time Surveillance for SARS-CoV-2 (VERSUS) Study: 11th Report. VERSUS Group. 2024 May 24 [Cited 2025 July 24]. Available from: https://www.tm.nagasaki-u.ac.jp/versus/results/20240524.html

4. Maeda H, Saito N, Igarashi A, Ishida M, Terada M, Masuda S, et al. Effectiveness of primary series, first, and second booster vaccination of monovalent mRNA COVID-19 vaccines against symptomatic SARS-CoV-2 infections and severe diseases during the SARS-CoV-2 omicron BA.5 epidemic in Japan: vaccine effectiveness real-time surveillance for SARS-CoV-2 (VERSUS). Expert Review of Vaccines. 2024;23(1):213–225. doi: 10.1080/14760584.2024.2310807. Epub 2024 Feb 8. PMID: 38288980.

5. Lazarus JV, Ratzan SC, Palayew A, Gostin LO, Larson HJ, Rabin K, et al. A global survey of potential acceptance of a COVID-19 vaccine. Nature Medicine. 2021;27(2):225–228. doi: 10.1038/s41591-020-1124-9. PMID: 33082575; PubMed Central PMCID: PMC7573523.

6. Fukunaga H. Social Psychology and Media Coverage of the New Corona Vaccination: An Internet Survey. The NHK Monthly Report on Broadcast Research. 2021;71(7):2–27. doi: 10.24634/bunken.71.7_2

7. Slovic P. Perception of risk. Science 1987;236(4799):280-285. doi: 10.1126/science.3563507. PMID: 3563507.

8. Romate J, Rajkumar E, Gopi A, Abraham J, Rages J, Lakshmi R, et al. What contributes to COVID-19 vaccine hesitancy?: A systematic review of the psychological factors associated with COVID-19 vaccine hesitancy. Vaccines (Basel) 2022;10(11):1777. doi: 10.3390/vaccines10111777. PMID: 36366286; PubMed Central PMCID: PMC9698528.

9. Freeman D, Loe BS, Chadwick A, Vaccari C, Waite F, Rosebrock L, et al. COVID-19 vaccine hesitancy in the UK: the Oxford coronavirus explanations, attitudes, and narratives survey (Oceans) II. Psychological Medicine 2022;52(14):3127–3141. doi: 10.1017/s0033291720005188. Epub 2020 Dec 11. PMID: 33305716; PubMed Central PMCID: PMC7804077.

10. Gregory ME, MacEwan SR, Powell JR, Volney J, Kurth JD, Kenah E, et al. The COVID-19 vaccine concerns scale: Development and validation of a new measure. Human Vaccines & Immunotherapeutics 2022;18(5):2050105. doi: 10.1080/21645515.2022.2050105. Epub 2022 Apr 5. PMID: 35380510; PubMed Central PMCID: PMC9196820.

11. Balgiu BA, Sfeatcu R, Dâncu AMC, Imre M, Petre A, Tribus L. The multidimensional vaccine hesitancy scale: A validation study. Vaccines (Basel) 2022;10(10):1755. doi: 10.3390/vaccines10101755. PMID: 36298620; PubMed Central PMCID: PMC9608997.

12. Wakashima K, Asai K, Kobayashi D, Koiwa K, Kamoshida S, Sakuraba M. The Japanese version of the Fear of COVID ― 19 scale: Reliability, validity, and relation to coping behavior. PLOS ONE 2020;15(11):e0241958. doi: 10.1371/journal.pone.0241958. PMID: 33152038; PubMed Central PMCID: PMC7644080.

13. Machida M, Kojima T, Popiel HA, Geiger M, Odagiri Y, Inoue S. Development, validity, and reliability of the Japanese version of the 7C of vaccination readiness scale. American Journal of Infection Control 2023;51(4):426–433. doi: 10.1016/j.ajic.2022.07.001. Epub 2022 Jul 15. PMID: 35839960.

14. Majima Y, Nakamura H. Development of the Japanese Version of the Generic Conspiracist Beliefs Scale (GCBS-J). Japanese Psychological Research 2019;62(4):254–267. doi: 10.1111/jpr.12267

15. Fukukawa Y, Oda R, Usami H, Kawahito J. Development of a Japanese version of the Perceived Vulnerability to Disease Scale. Japanese Journal of Psychology. 2014;85(2):188–95. doi: 10.4992/jjpsy.85.13206

16. Masuda S, Sakagami T, Morii M. Comparison among methods for improving response quality of surveys. Japanese Journal of Psychology. 2019;90(5):463–472. doi: 10.4992/jjpsy.90.18042

17. Al Baseer NAM, Shaheen HS. A study of some psychological variables as predictors of COVID-19 vaccination anxiety among Faculty Members of Ain Shams University. Scientific Reports. 2024;14:26615. doi: 10.1038/s41598-024-75360-x

18. Lee J, Paek I. In search of the optimal number of response categories in a rating scale. Journal of Psychoeducational Assessment. 2014;32:663¬673. doi: 10.1177/0734282914522200

19. Lozano LM, García-Cueto E, Muñiz J. Effect of the number of response categories on the reliability and validity of rating scales. Methodology: European Journal of Research Methods for the Behavioral and Social Sciences. 2008;4(2):73¬79. doi: 10.1027/1614-2241.4.2.73

20. Sekizawa Y, Hashimoto S, Denda K, Ochi S, So M. Association between COVID-19 vaccine hesitancy and generalized trust, depression, generalized anxiety, and fear of COVID-19. BMC Public Health. 2022;22(1):126. doi: 10.1186/s12889-021-12479-w. PMID: 35042506; PubMed Central PMCID: PMC8764499.

21. Al-Qazaz HK, Al-Obaidy LM, Attash HM. COVID-19 vaccination, do women suffer from more side effects than men? A retrospective cross-sectional study. Pharm Pract (Granada). 2022;20(2):2678. doi: 10.18549/PharmPract.2022.2.2678. Epub 2022 Jun 10. PMID: 35919795; PubMed Central PMCID: PMC9296083.

22. Lewis A, Duch R. Gender differences in perceived risk of COVID-19. Soc Sci Q. 2021;102(5):2124–2133. doi: 10.1111/ssqu.13079. Epub 2021 Oct 29. PMID: 34908608; PubMed Central PMCID: PMC8662048.

23. Duncan LA, Schaller M, Park JH. Perceived vulnerability to disease: Development and validation of a 15-item self-report instrument. Personality and Individual Differences. 2009;47:541–546. doi: 10.1016/j.paid.2009.05.001

24. Lee SK, Sun J, Jang S, Connelly S. Misinformation of COVID-19 vaccines and vaccine hesitancy. Scientific Reports. 2022;12(1):13681. doi: 10.1038/s41598-022-17430-6. PMID: 35953500; PubMed Central PMCID: PMC9366757.

